# The Association of the Microbiome with Melanoma Tumor Response to Immune Checkpoint Inhibitor Treatment and Immune-Related Adverse Events (NCT05102773)

**DOI:** 10.1101/2025.01.30.25321413

**Authors:** Caroline Dravillas, Nyelia Williams, Marium Husain, Rebecca Hoyd, Ahmed Hussein, Alexa Meara, Mari Lynn, Amna Bibi, Bailey Conrad, Noah Lepola, Shannon Gray, Michael Bodnar, Namrata Arya, Scott Roberts, Phuong Hoang, Jessica Apparicio, Deanna Merrill, Richard Wu, Claire Verschraegen, Christin E. Burd, Kari Kendra, Dan Spakowicz

**Author notes:** These authors contributed equally (Caroline Dravillas, Nyelia Williams).

## Abstract

Improved understanding of the factors that underlie immune checkpoint inhibitor (ICI) response and toxicity are needed as only half of patients with metastatic melanoma respond, and 10-40% experience immune-related adverse events (irAEs). Modifying the gut microbiome could positively affect response to ICIs and reduce toxicities. Here, we sought to determine if the pre-treatment gut microbiome predicts ICI response or toxicity in the setting of metastatic melanoma. Melanoma patients (n=88) over 18 years of age, planning to receive ICI therapy enrolled in a prospective observational cohort study at The Ohio State University Comprehensive Cancer Center Skin Cancer Clinic. Patients taking corticosteroids for indications other than adrenal physiologic replacement were excluded. Stools were collected at baseline, within 10 days of an irAE as determined by CTCAE v 5.0 criteria, and at 12 weeks. ICI response and progression-free survival (PFS) were evaluated q12 weeks using Response Evaluation Criteria in Solid Tumors (RECIST v1.1). Metagenomic whole-genome shotgun sequencing of the microbiome was classified using MetaPhlAn4/HUMAnN3 and differential abundance analyzed with ANCOM-BC2. Of the 88 patients enrolled, 41 had metastatic disease and complete data. There were 25 participants classified as responders, defined as having complete response or partial response according to RECIST criteria, or stable disease with 6-month PFS. Grade ≥ 1 irAEs were observed in 15/41 participants. The abundance of *Intestinimonas butyriciproducens* (q-value = 0.002) and *Longicatena caecimuris* (q-value = 0.003) were enriched in responders, *Tenericutes* (q-value= 0.001) and *Lachnospira sp. NSJ 43* (q-value =0.002) in non-responders. *Blautia luti*, as well as several other *Lachnospiraceae*, were associated with response and no irAE (response q-value = 0.02, no irAE q-value = 0.02). The association of response to ICIs with several taxa in the family *Lachnospiraceae*, a prevalent microbial family in the gut, is consistent with prior research, which has found that this family may influence treatment outcomes through various mechanisms, such as immune regulation, metabolism, and pathogen exclusion. While no statistical relationship was observed between response and irAEs in this cohort, the microbes associated with both could serve as biomarkers. Future studies to assign causal roles for (specific microbes) in response and toxicity could identify mechanisms to improve patient outcomes.

## Introduction

Immune checkpoint inhibitor (ICI)-based immunotherapy is the standard of care for multiple cancer types. ICIs trigger immune-related adverse events (irAEs) in 40% of patients.[1] Among those experiencing an irAE, half (20%) will experience symptoms that affect their quality of life and necessitate chronic treatment.[1] irAEs can impact any organ, and in very rare cases, a fatal outcome can occur after only a single immunotherapy dose. Our understanding of irAE pathophysiology is fragmented, and treatment decisions to avoid or address irAEs are being made with limited scientific evidence.

The gut microbiome is associated with ICI response and irAEs.[2–6] Early studies reported decreased survival in individuals receiving antibiotics around the time of ICI initiation.[7,8] This finding led to prospective trials in which microbes in the gut were found to correlate with clinical outcomes in mouse studies.[9] Further, fecal transplantation of a responder or healthy donor’s gut microbiome into patients who progressed on ICIs resensitized the patient to the same ICI treatment.[10,11] Despite these observations, the microbes and mechanisms by which they impact ICI outcomes is not fully understood. Additionally, individuals who develop irAEs tend to show improved treatment response if the symptoms can be effectively managed.[12–14] The microbiome holds promise to prevent irAEs and improve ICI treatment response.

Here, we describe the results of a prospective, longitudinal study of gut microbiome samples from melanoma patients treated with ICIs (Clinicaltrials.gov: NCT05102773). The trial aimed to determine if the microbiome α-diversity predicts response per Response Evaluation Criteria in Solid Tumors (RECIST) v1.1 at a 12-week computed tomography (CT) scan or toxicity per CTCAEv5.0. Secondarily, it aimed to determine the recruitment and compliance rates for longitudinal biospecimen collection, including stool, in melanoma patients. An exploratory objective included determining if individual microbes or their changes in relative abundance are predictive of response or toxicity. We show how a stool sample collected within 10 days of treatment initiation can predict treatment response and irAE development. We highlight microbes associated with an improved response but no irAE development.

## Methods

### Patient Cohort

This prospective observational cohort study was approved by the Ohio State University Comprehensive Cancer Center (OSUCCC) Institutional Review Board (protocol 2019C0151) and registered as NCT05102773. Patients were recruited from the OSUCCC Skin Cancer Clinic. Patients were excluded if they were on corticosteroids for any reason other than adrenal physiologic replacement therapy at the start of the first ICI cycle. Stool samples were collected at baseline (before ICI started), within 10 days of an irAE, and at 12 weeks. Blood samples were also collected at baseline. The patient cohort was filtered to include only those with metastatic disease, response information, and a baseline microbiome sample.

### Clinical Outcome Assessment

Response to ICIs was evaluated at 12 weeks using the Response Evaluation Criteria in Solid Tumors (RECIST v1.1).[15] Patients were classified into RECIST groups: complete response (CR), partial response (PR), stable disease (SD), and progressive disease (PD). Progression-free survival (PFS) was also recorded. Responders were defined as having complete response, partial response, or stable disease with 6-month PFS. Those with stable disease lacking 6-month PFS and those with progressive disease were classified as non-responders. Adverse events were evaluated using the Common Terminology Criteria for Adverse Events (CTCAE v5.0).[16]

### Data generation, processing, and analysis

Metagenomic whole-genome shotgun (WGS) sequencing was performed on the Illumina NovaSeq 6000 platform. Before taxonomic classification, FastQC was used to assess the quality of WGS reads.[17] It was determined that no further quality control or trimming was necessary. Taxonomic classification and profiling were performed using MetaPhlAn4 and HUMAnN3.[18,19] Descriptive statistics were used to summarize the cohort. For continuous variables, comparisons between the groups were made using Student’s t-tests. For categorical variables, group comparisons were made using the chi-squared or Fisher’s exact tests. Differential abundance analysis was performed using ANCOM-BC2 (Analysis of Composition of Microbiomes with Bias Correction 2) to identify significant taxonomic differences between responders and non-responders at baseline.[20] Structural zeroes, microbes absent in at least 1 group, were also identified during this process. The same analysis was performed to asses differences in the baseline microbial composition of those who did and did not experience an irAE. Results were considered significant at a q-value of <0.05. All statistical analyses were performed in R.

### Data availability

Processed relative abundance and count tables, along with the code to regenerate all analyses and figures, are available at https://github.com/spakowiczlab/mitox.

## Results

We assessed patients from the OSUCCC Skin Clinic who were scheduled to receive ICIs for stage IV melanoma for study eligibility. We approached 112 patients about participation, of which 88 consented (79%). Of these patients, 42 completed the study (48%), including collecting at least 1 gut microbiome sample (**Figure 1A**). The study lasted 16 weeks; patients were asked to provide blood and gut microbiome samples at baseline (within 10 days of starting treatment), 12 weeks after beginning treatment, and upon developing any grade irAE (**Figure 1B**). There was some variability in the timing of study events, with all but 1 participant collecting a baseline gut microbiome sample within 10 days of their cycle 1 day 1 (C1D1) treatment visit (**Figure 1C**).

**Figure 1.**
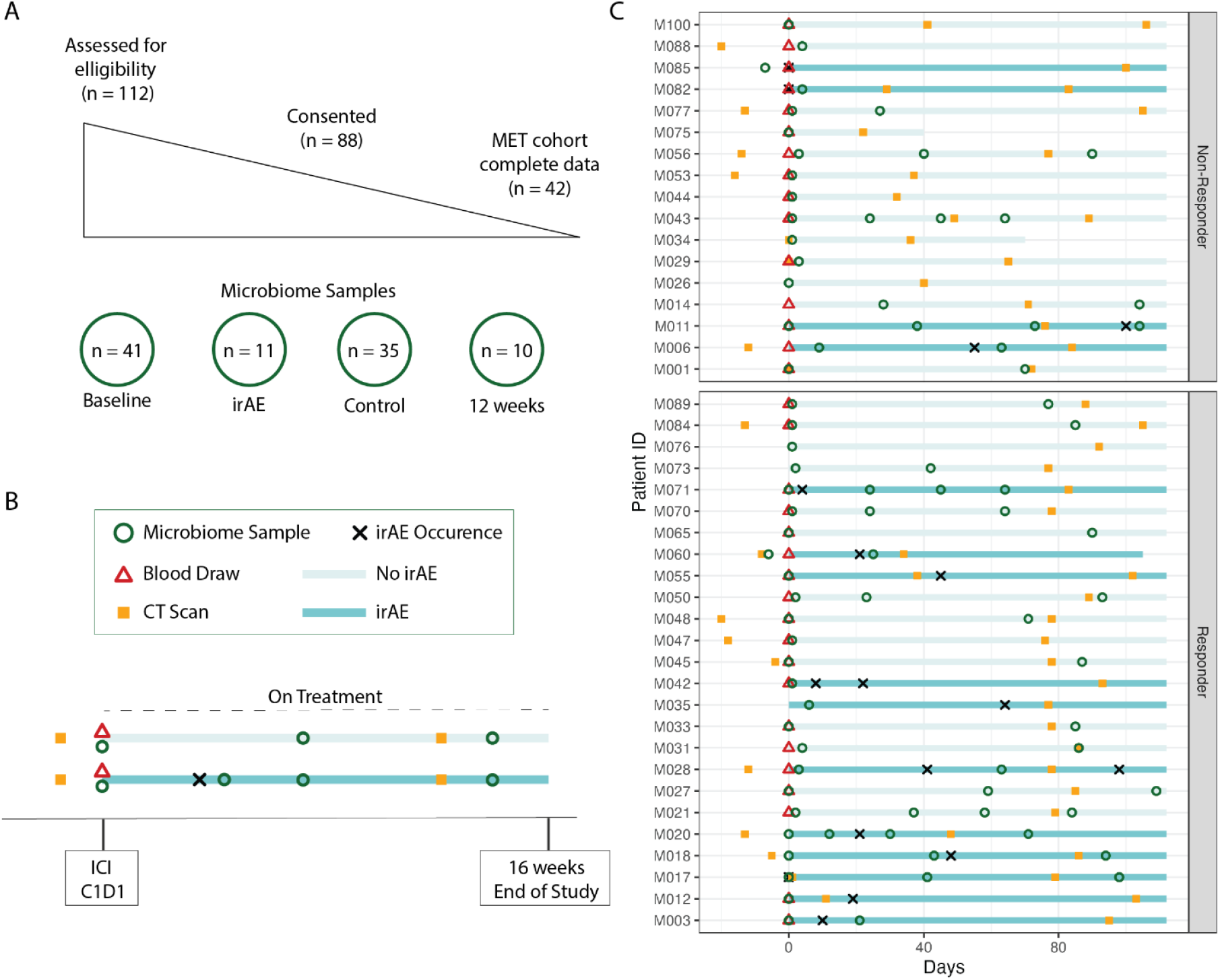
Summary of the study timeline and sample collection. **A)** Visualization of participant recruitment, consent, inclusion, and sample collection showing the final cohort, and the number of samples collected at each time point. **B)** Study schema depicting trial timeline. Patients with metastatic melanoma were asked to collect blood and microbiome samples within 10 days of ICI cycle 1 day 1 (C1D1), at their 12-week computed tomography (CT) scan, and in the event of any grade irAE within 16 weeks of C1D1. **C)** A timeline for each patient, showing the variation in microbiome sample collection, CT scan timing, response, and irAE development.

Treatment response was categorized according to RECIST v1.1 at the 12-week CT scan; those with complete response, partial response, and stable disease ≥ 6 months were classified as responders (R, *n*=25), and those with progression or stable disease for < 6 months were classified as non-responders (NR, *n*=16) (**Table 1**). In this study, 60% of tumors responded to ICI treatment. There was no difference in age, sex, race, treatment type, or stage between the R and NR groups. Further, we saw no correlation between irAE development and treatment response (*P*=0.368).

**Table 1.**
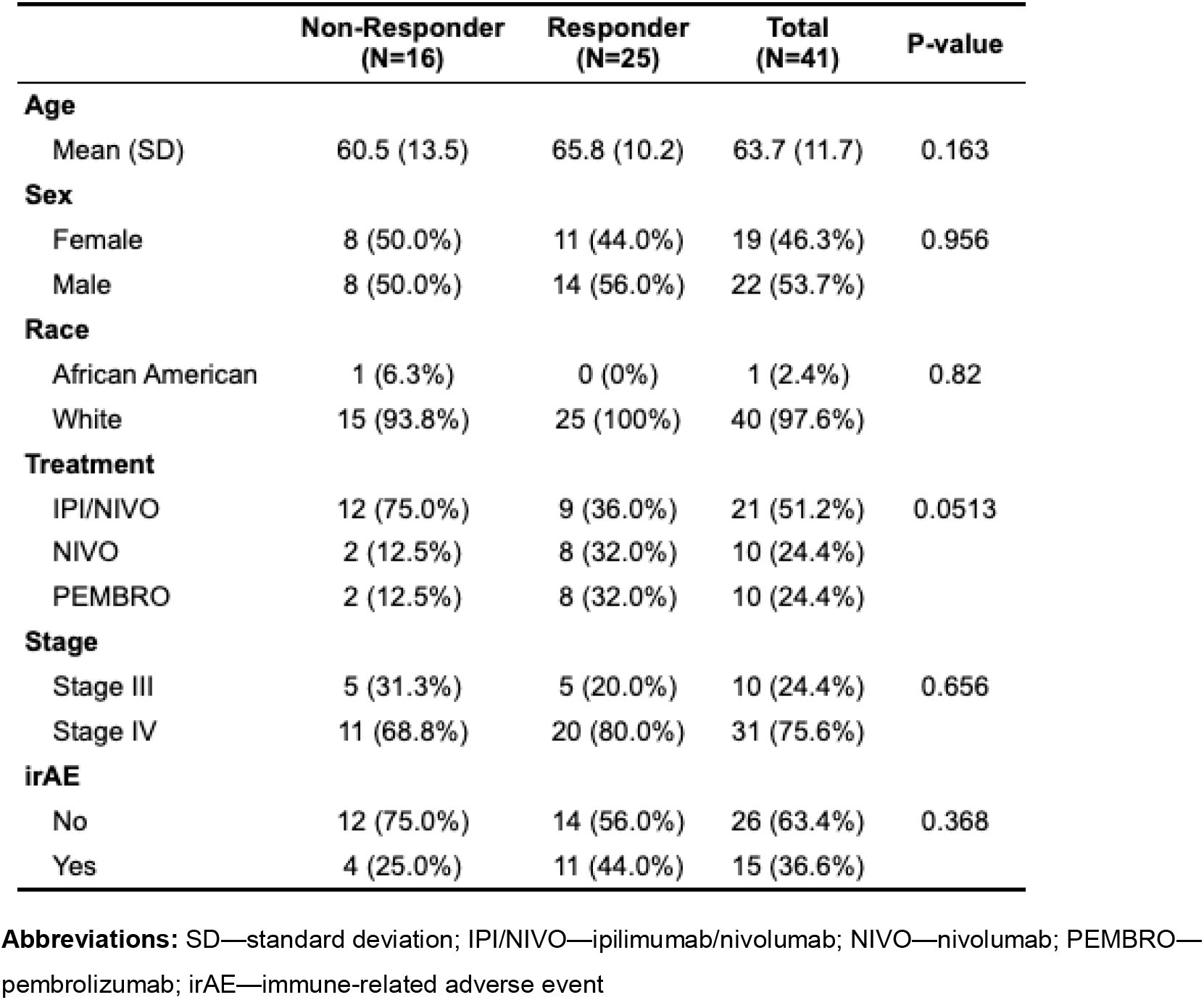
Cohort demographics stratified by treatment response.

|  | Non-Responder<br>(N=16) | Responder<br>(N=25) | Total<br>(N=41) | P-value |
| --- | --- | --- | --- | --- |
| **Age** |  |  |  |  |
| Mean (SD) | 60.5 (13.5) | 65.8 (10.2) | 63.7 (11.7) | 0.163 |
| **Sex** |  |  |  |  |
| Female | 8 (50.0%) | 11 (44.0%) | 19 (46.3%) | 0.956 |
| Male | 8 (50.0%) | 14 (56.0%) | 22 (53.7%) |  |
| **Race** |  |  |  |  |
| African American | 1 (6.3%) | 0 (0%) | 1 (2.4%) | 0.82 |
| White | 15 (93.8%) | 25 (100%) | 40 (97.6%) |  |
| **Treatment** |  |  |  |  |
| IPI/NIVO | 12 (75.0%) | 9 (36.0%) | 21 (51.2%) | 0.0513 |
| NIVO | 2 (12.5%) | 8 (32.0%) | 10 (24.4%) |  |
| PEMBRO | 2 (12.5%) | 8 (32.0%) | 10 (24.4%) |  |
| **Stage** |  |  |  |  |
| Stage III | 5 (31.3%) | 5 (20.0%) | 10 (24.4%) | 0.656 |
| Stage IV | 11 (68.8%) | 20 (80.0%) | 31 (75.6%) |  |
| **irAE** |  |  |  |  |
| No | 12 (75.0%) | 14 (56.0%) | 26 (63.4%) | 0.368 |
| Yes | 4 (25.0%) | 11 (44.0%) | 15 (36.6%) |  |

**Abbreviations:** SD—standard deviation; IPI/NIVO—ipilimumab/nivolumab; NIVO—nivolumab; PEMBRO—pembrolizumab; irAE—immune-related adverse event

The baseline microbiomes of both the R and NR groups were dominated by *Firmicutes* and *Bacteroidota* **(Figure 2A**). However, patients in the R group had more *Lachnospiraceae*, including several *Blautia* and *Enterocloster* species (**Figure 2B**), while those in the NR group were enriched for *Tenericutes*. Microbes only observed in 1 category included *Enterobacter* in the NR group and *Bacteroides salyersiae* in the R group (**Figure 2C**).

**Figure 2.**
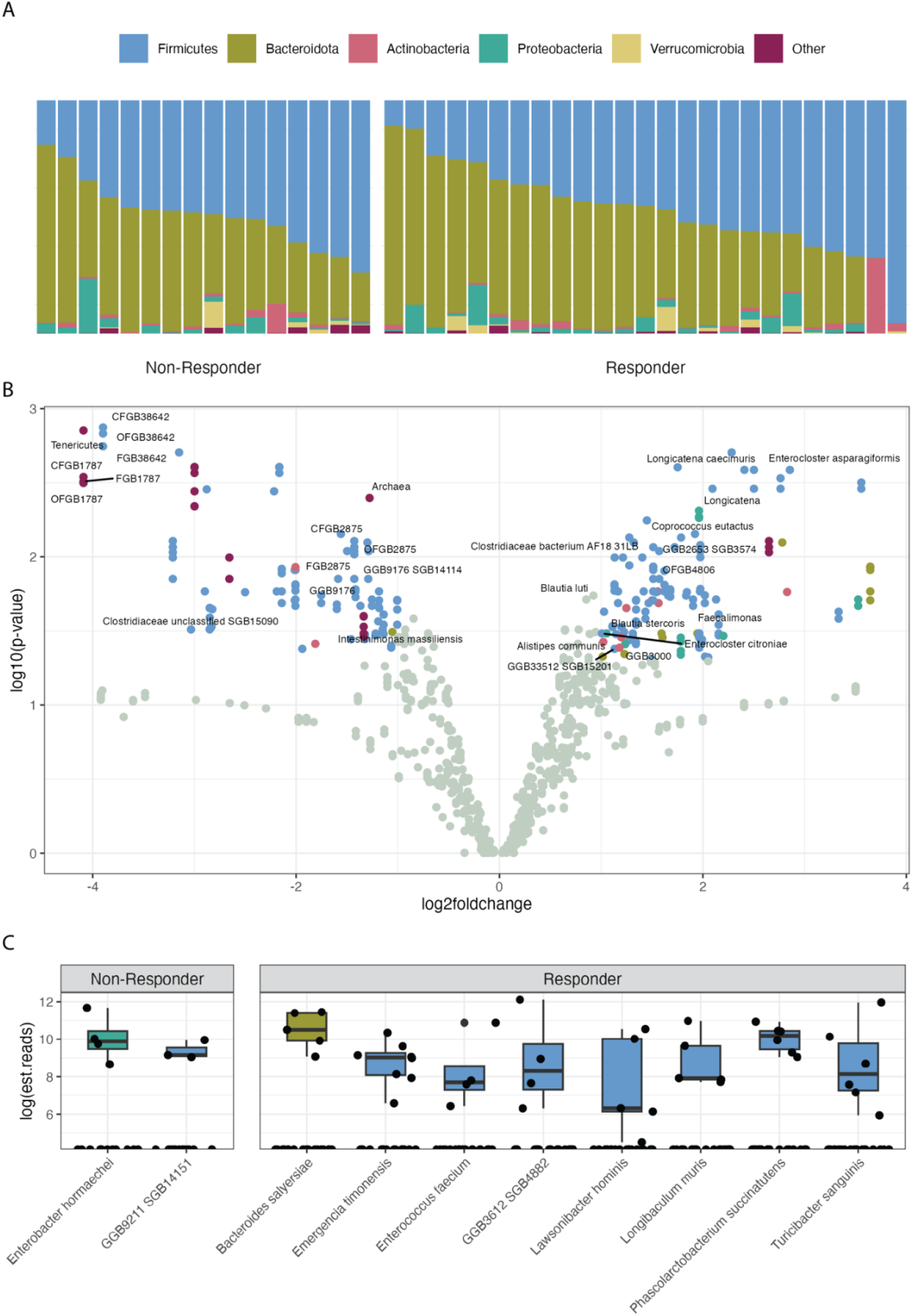
Differential abundance of baseline gut microbes between R and NR. **A)** Relative abundance of the dominant phyla in each patient. **B)** Enrichment of individual microbes, at all taxonomic levels, in R (right) vs. NR (left) with the false discovery rate-corrected *P*-values. **C)** Microbes found only in 1 category or another and present in at least 15% of the samples in that category.

Individuals who developed an irAE did not differ in age, sex, race, treatment type, stage, or treatment response (**Table 2**).

**Table 2.** Cohort demographics stratified by the development of an irAE.

|  | irAE<br>(N=15) | No irAE<br>(N=26) | Total<br>(N=41) | P-value |
| --- | --- | --- | --- | --- |
| **Age** |  |  |  |  |
| Mean (SD) | 60.8 (13.3) | 65.4 (10.6) | 63.7 (11.7) | 0.232 |
| **Sex** |  |  |  |  |
| Female | 9 (60.0%) | 10 (38.5%) | 19 (46.3%) | 0.314 |
| Male | 6 (40.0%) | 16 (61.5%) | 22 (53.7%) |  |
| **Race** |  |  |  |  |
| White | 15 (100%) | 25 (96.2%) | 40 (97.6%) | 1 |
| African American | 0 (0%) | 1 (3.8%) | 1 (2.4%) |  |
| **Treatment** |  |  |  |  |
| IPI/NIVO | 11 (73.3%) | 10 (38.5%) | 21 (51.2%) | 0.0642 |
| NIVO | 1 (6.7%) | 9 (34.6%) | 10 (24.4%) |  |
| PEMBRO | 3 (20.0%) | 7 (26.9%) | 10 (24.4%) |  |
| **Stage** |  |  |  |  |
| Stage III | 3 (20.0%) | 7 (26.9%) | 10 (24.4%) | 0.905 |
| Stage IV | 12 (80.0%) | 19 (73.1%) | 31 (75.6%) |  |
| **Response** |  |  |  |  |
| Non-Responder | 4 (26.7%) | 12 (46.2%) | 16 (39.0%) | 0.368 |
| Responder | 11 (73.3%) | 14 (53.8%) | 25 (61.0%) |  |

**Abbreviations:** irAE—immune-related adverse event; SD—standard deviation; IPI/NIVO—ipilimumab/nivolumab; NIVO—nivolumab; PEMBRO—pembrolizumab

We observed no phylum-level differences between those who developed an irAE and those who did not (**Figure 3A**). Regarding individual microbes, the no-irAE group had more *Lachnospiraceae*, including several *Anaerotignum* and *Enterocloster* species (**Figure 3B**), while the irAE group contained more *Bifidobacterium*. Microbes only observed in 1 category included a metagenome-assembled taxon in the family *Oscillospiraceae* in the no-irAE group, and *Bifidobacterium dentium* in the irAE group (**Figure 3C**).

**Figure 3.**
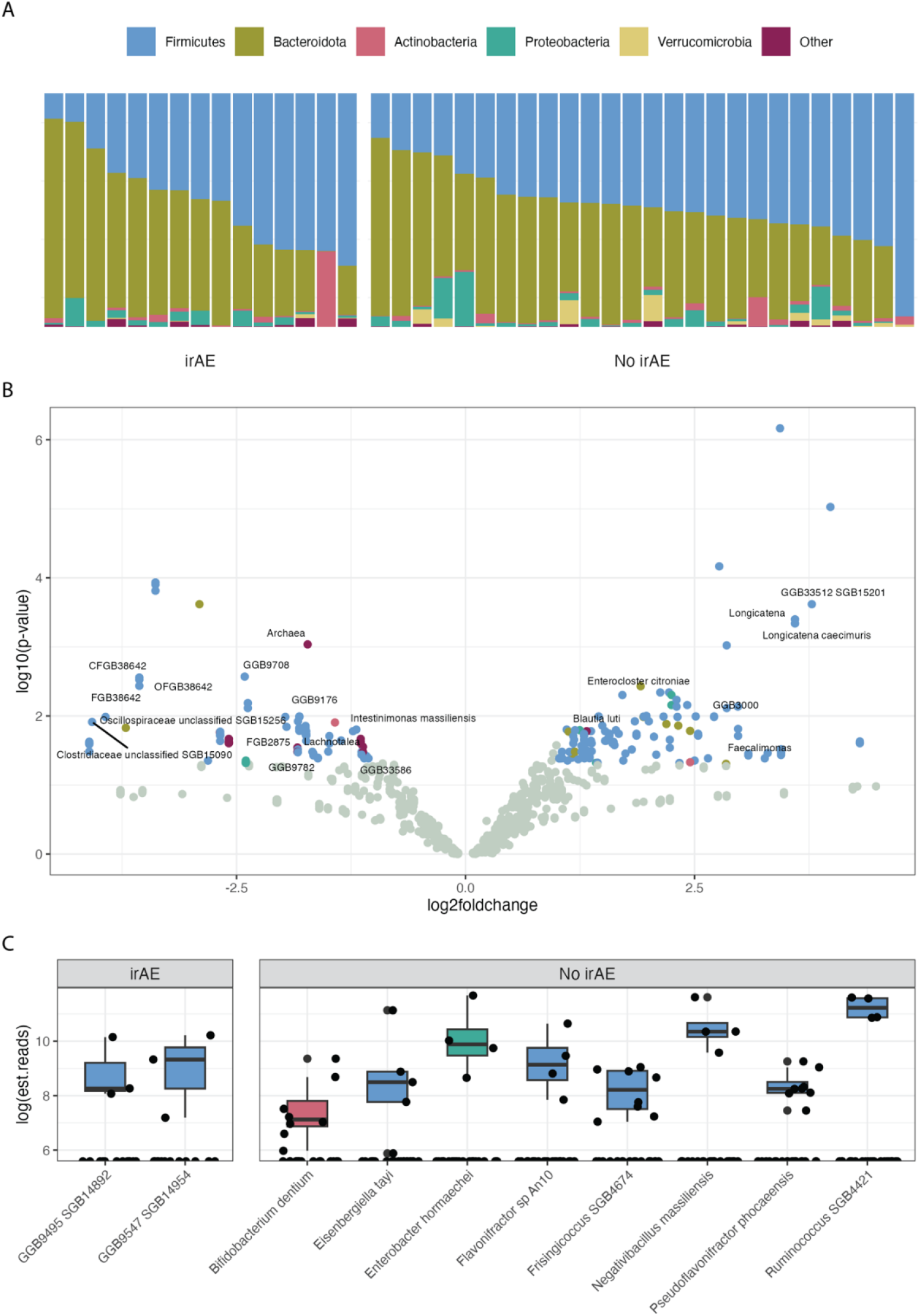
Differential abundance of baseline gut microbes in the irAE and no-irAE groups. **A)** Relative abundance of the dominant phyla in each patient. **B)** Enrichment of individual microbes, at all taxonomic levels, in the no-irAE (right) versus irAE (left) groups with the false discovery rate corrected *P*-values. **C)** Microbes found in a single group (no-irAE or irAE) and present in at least 15% of the samples in that category.

## Conclusions

The clinical trial NCT05102773, “The effect of the microbiome on immune checkpoint inhibitor toxicity in patients with melanoma,” studied the microbiomes of 42 patients undergoing ICI treatment for metastatic melanoma. At baseline, many microbes stratified the cohort by treatment response according to RECIST criteria, and a smaller number of microbes were associated with individuals who developed an irAE. The sets of microbes associated with response and irAE development were partially overlapping—i.e., some microbes were enriched in patients with a positive response to treatment who did not develop an irAE—suggesting these 2 variables could be deconvoluted with careful curation of the microbiome. This observation suggests that although irAE events and response have been correlated in the past, it may be possible to modify the microbiome in ways that would enhance treatment response while decreasing the risk of irAEs.

The microbes associated with the R and NR groups were largely consistent with the literature, with a few intriguing exceptions. We observed many family members of *Lachnospiraceae* in the taxa associated with responders. A recent meta-analysis of 5 studies found *Lachnospiraceae* to be the most consistently associated with ICI response.[21,22] Further, *Lachnospiraceae* is the most prevalent gut microbial family across human populations worldwide and can reach up to 20% of the healthy gut microbiome.[23] *Lachnospiraceae* contributes to metabolism (of dietary fiber, xenobiotics, and host compounds like bile acids), immune regulation (via producing short-chain fatty acids and degrading lyso-glycerophospholipids), and niche exclusion (of, for example, *Clostridioides difficile* and vancomycin-resistant *Enterococcus*). However, *Lachnospiraceae* are genetically diverse, with large pan-genomes, suggesting more host interactions to be found. The microbiome has been associated with ICI response via several mechanisms, including antigen mimicry [24], innate immune activation (for example, via toll-like receptors [TLRs]) [25], and small-molecule secretion distal to [26] and within [27] the tumor. Therefore, the associations presented here likely represent the activity of several independent mechanisms. Notably, we observed several *Blautia* spp. significantly associated with R, which were recently associated with improved ICI response in model systems and for which Jahanbakhshi et al. proposed a carnitine-dependent mechanism.[28]

Many microbes associated with R were also associated with the development of an irAE. However, we observed no statistical relationship between these outcomes in this study, even though irAEs have been associated with longer survival in larger cohorts.[29,30] Some irAEs are easily managed, for example, irAE-thyroiditis can be addressed by thyroid replacement therapy.[29,31] However, toxicities such as pneumonitis [32,33], cytopenia [34], encephalitis, hepatitis, myocarditis [35], and severe irAEs occurring in older patients [12] are associated with worse survival. Despite little evidence to support and guide steroid dose or duration, the most general treatment strategy is high-dose administration. There is a lack of understanding of how steroids and other immunosuppressive therapies may impair antitumor immunity, and there is potential for long-term side effects. We and others have previously studied potential clinical risk factors and predictive biomarkers for irAEs [12,32,36]. Still, prospective studies have not validated these or provided enough mechanistic insights to inform therapeutic interventions. Therefore, there remains a clinical need to develop therapies to specifically address irAEs, particularly as new biologics used to treat autoimmune diseases have not been widely explored in the context of cancer. The microbes associated with both R and irAE (e.g., *Enterocloster citroniae*) may provide a biomarker for irAE risk and the microbes associated with R and no-irAE (e.g., *Blautia luti*) a therapeutic target for microbiome modulation.

Limitations of this study include the relatively small sample size (*n*=42 assessable for response), particularly with respect to irAE development (*n*=11 developed a grade >1 irAE), which may limit the generalizability of the findings. Patients on corticosteroids at the start of ICI cycle 1 were excluded, which may introduce selection bias. The study design does not establish causality between the microbiome and either of the explored patient outcomes. There was no effort to control for lifestyle factors known to affect the microbiome, such as diet. The sample collection timing was relatively broad (a window of 10 days), and the 3 sample time points are not expected to capture the full dynamics of microbiome changes during ICI administration. Finally, the study was conducted at a single center (OSUCCC), with a narrow demographic range, which may limit the generalizability of the findings to other populations or clinical settings.

In summary, we identified baseline microbiomes associated with response and toxicity in our study of 41 patients undergoing treatment with ICIs for metastatic melanoma. This suggests a potential biomarker for treatment outcomes and treatment-related toxicity, as well as a therapeutic target to improve patient outcomes. Additional work is needed to identify the causal relationships among these associations, define the mechanisms eliciting these effects, and devise an appropriate strategy to leverage this knowledge to improve health.

## Data Availability

Data and code to reproduce all analyses and figures is available at https://github.com/spakowiczlab/mitox.

## Funding

Pelotonia, clinical trial award (DS)

National Institutes of Aging grant 1K01AG070310 (DS)

The Ohio State University Wexner Medical Center and The Ohio State University College of Medicine Clinical Research Center/Center for Clinical Research Management

## Abbreviations

CT: computed tomography
CTCAE: Common Terminology Criteria for Adverse Events
ICI: immune checkpoint inhibitor
IPI/NIVO: ipilimumab/nivolumab
irAE: immune-related adverse event
NIVO: nivolumab
NR: non-responder
OSUCCC: The Ohio State University Comprehensive Cancer Center
PEMBRO: pembrolizumab
PFS: progression-free survival
R: responder
RECIST: Response Evaluation Criteria in Solid Tumors
SD: standard deviation

